# Modeling the Future Incidence of Preeclampsia under Climate Change and Population Growth Scenarios

**DOI:** 10.1101/2024.12.20.24319323

**Authors:** Iaroslav Youssim, Daniel Nevo, Offer Erez, Chaim I. Garfinkel, Barbara S. Okun, Lena Novack, Itai Kloog, Raanan Raz

## Abstract

Preeclampsia is a dangerous pregnancy disorder, with evidence suggesting that high ambient temperatures may increase its risk, making future incidence projections crucial for health planning. While temperature-related projections for all-cause mortality exist, disease-specific projections, especially for pregnancy complications, are limited due to data and methodological challenges. Vicedo-Cabrera et al. (2019) pioneered a time-series approach to project health impacts using the attributable fraction (AF) of cases due to climate change. We adjusted this method for preeclampsia, whose risk involves long-term exposures, with delivery as a competing event.

We based our analysis on the exposure-response relationship estimated in our previous study in southern Israel using cause-specific hazard and distributed lag nonlinear models. In the current study, we modeled several demographic and climate scenarios in the region for 2020-2039 and 2040-2059. Scenario-specific AFs were calculated by comparing cumulative preeclampsia incidence with and without corresponding climate change. Finally, annual cases were estimated by applying climate scenario-specific AFs to cases projected under each demographic scenario.

Our models show that climate change alone may increase preeclampsia by 3.2% to 4.3% in 2040-2059 relative to 2000-2019. Fertility trends are modeled to have a larger impact, with a 30% increase in cases by 2020-2039 under a low-fertility scenario. Extreme high-fertility and climate scenarios could result in a 2.3-fold rise in incidence, from 486 cases annually in 2000-2019 to 1,118 by 2040-2059.

## 1. Introduction

Preeclampsia is a dangerous pregnancy disorder characterized by new-onset hypertension combined with proteinuria, accounting for an annual global toll of half a million fetal and 70,000 maternal deaths and other complications (Erez et al., 2022; Jung et al., 2022; Rana et al., 2019). The expected incidence of preeclampsia is an important factor for planning community and hospital services that supply prevention, management, and treatment for preeclampsia and its complications.

Several recent epidemiological investigations from different countries demonstrated associations between high prenatal ambient temperature and increased risk of preeclampsia (Part et al., 2022; Shashar et al., 2020; Xiong et al., 2020; Youssim et al., 2023), suggesting that climate change may increase its incidence. As expected, the exposure-response relations for temperature and preeclampsia vary by study. Such variation can result from different study designs and statistical methods but is also expected due to differences in climatic zones, temperature ranges, and populations. In addition, the absolute number of incident cases is also affected by the number of births in a population, which is impacted by changes in fertility rates.

Vicedo-Cabrera et al. suggest a modeling framework for projections of climate change impacts on health (Vicedo-Cabrera et al., 2019), composed of four phases: 1) Assessing the exposure-response function based on historical data; 2) Evaluating possible changes in the exposure distribution using climate models; 3) Estimating the baseline rate of the health outcome in the future using demographic projections; and 4) Combining these factors to model future incidence under each scenario, using the epidemiological measure of attributable fraction (AF). This enables the modeling of the number or percentage of cases attributed to climate change. In recent years, led by this general framework, time-series analyses were employed with distributed-lag Poisson models attempting to project the future burden of temperature-related all-cause mortality (Gu et al., 2020). Yet similar projection studies of specific diseases are still rare (Onozuka et al., 2019). Since most existing temperature and health epidemiological studies investigate the short-term effects of acute outcomes, they typically use time-series analyses, as with the methodology described above (Vicedo-Cabrera et al., 2019). Preeclampsia risk, however, can be impacted by a longer-term exposure, such as during the first pregnancy trimester, and may take more time to develop (Erez et al., 2022). In addition, birth is a competing event for preeclampsia since birth without the disorder prevents its development (Reeder et al., 2023). Therefore, the exposure-response relations between temperature and preeclampsia are better modeled using cause-specific survival models with distributed lag nonlinear models (DLNMs). This requires a modification of the methodology proposed by Vicedo-Cabrera and colleagues, in which AF is quantified by comparing the cumulative incidence expected with and without a defined change in the exposure distribution, compliant with a specific climate scenario.

Many previous studies projecting health effects assumed constant future populations (Cole et al., 2023; Vicedo-Cabrera et al., 2019). However, this is not the case in the Israeli population, which demonstrates strikingly high levels of population growth rates and fertility, especially compared with comparable countries in terms of their education and income. Israel’s population growth rate stood near 1.9%, and period total fertility stood around 3.0 in 2021 (Israel Central Bureau of Statistics, 2023). The current growth rate implies a population doubling time of roughly 35 years. Moreover, in recent decades, most of the population growth has been due to natural increases, although in the past, migration was a major source of growth within the Jewish population. The population of Israel is also extremely heterogeneous, with differing family formation patterns across main social groups. There are substantial differences in demographic patterns between the minority Arab population (primarily Sunni Muslim) and the majority Jewish population. Moreover, there is great heterogeneity in demographic patterns within population groups, particularly by levels of religiosity (Okun, 2013). Therefore, population projections need to account for the heterogeneity in the population in terms of demographic patterns.

In this study, we estimate possible scenarios of change in preeclampsia incidence in southern Israel during two 20-year future intervals, 2020-2039 and 2040-2059, by adjusting the general methodology proposed by Vicedo-Cabrera and colleagues. We rely on our previous work estimating the historical associations between temperature and preeclampsia, using data on pregnancies from 2005 to 2019 at Soroka University Medical Center in Be’er Sheva, Israel (Youssim et al., 2023). We further model expected temperature exposure and fertility rates in the geographic area served by this medical center. Finally, we estimate the separate and combined effects of fertility rates and climate change scenarios on the incidence of preeclampsia.

## 2. Methods

### 2.1 Study design and study population

The estimation of historical associations between prenatal exposure to ambient temperature and preeclampsia is based on our previous publication (Youssim et al., 2023). Briefly, we used a historical cohort design with time-to-event data on 128,377 live births from 2005 to 2019 at Soroka University Medical Center in Beer Sheva, Israel (henceforth Soroka). The births were spatially joined with satellite-based ambient temperature models using residential addresses, dates of birth, and gestational age at birth. Cause-specific hazard models with distributed lag functions were used to estimate the associations between temperature exposure during pregnancy and preeclampsia. In the current analysis, we used these exposure-response curves and additional data from climate and demographic projections to model the future burden of preeclampsia under diverse temperature and fertility scenarios. This modeling approach assumes the underlying associations’ causality and stability over time and does not take into account possible physiological or technological adaptation to climate changes.

### 2.2 Demographic projections

Projections of annual numbers of births by ethnicity (Jewish/Arab) for the population in the Be’er Sheva subdistrict for 2022-2059 were modeled under three scenarios: low, medium, and high fertility, based on population forecasts made by the Israel Central Bureau of Statistics (ICBS). The detailed methodology and assumptions of the population forecasts can be found in Israel Population Forecasts 2015-2065 (Faran and Klinger, 2018). Separate scenarios were applied for non-ultraorthodox Jews, ultraorthodox Jews, and Arabs because the fertility rates and population growth rates differ dramatically across these groups. For all groups, mortality was assumed to follow the medium-level projection, and migration was assumed to be negligible. The three fertility scenarios are based on ICBS assumptions for the fertility of each group for the entire country for the year 2065, and the initial fertility rates for each group are assumed to change linearly until that year. The age patterns of fertility in each group were assumed to be constant over the period.

Historical numbers of births in 2000-2021 in the Be’er Sheva subdistrict were also extracted from the ICBS (Israel Central Bureau of Statistics, 2022). Population growth coefficients projected under each fertility scenario were calculated for each 20-year interval (2020-2039 and 2040-2059) as the ratio of the projected to the historical number of births, where the historical interval was defined as 2000-2019. The expected number of births at Soroka for each interval was modeled as a product of the population growth coefficients with the total number of births observed in Soroka from 2000 to 2019. Next, the number of preeclampsia cases during each future 20-year time interval was modeled as a function of the fertility scenarios by multiplying the projected numbers of births at Soroka with the incidence observed there from 2005 to 2019 (Table 1).

**Table 1.** Historical and projected numbers of births in Beer Sheva subdistrict and in Soroka Medical Center under three demographic scenarios assuming medium mortality

| Scenario | Time interval | Population growth coefficient,<br><sup>a</sup> | N births, Soroka, <sup>b</sup> | Cases of preeclampsia, <sup>c</sup> | Preeclamp sia/annum |
| --- | --- | --- | --- | --- | --- |
| Historical | 2000-2019 | 1.0 | 247,445 | 9,718 | 486 |
| Projected |  |  |  |  |  |
| Low fertility | 2020-2039 | 1.33 | 329,186 | 12,929 | 646 |
|  | 2040-2059 | 1.37 | 339,857 | 13,348 | 667 |
| Medium fertility | 2020-2039 | 1.43 | 352,521 | 13,845 | 692 |
|  | 2040-2059 | 1.80 | 445,578 | 17,500 | 875 |
| High fertility | 2020-2039 | 1.50 | 371,371 | 14,586 | 729 |
|  | 2040-2059 | 2.21 | 545,636 | 21,430 | 1,071 |

<sup>a</sup> Population growth coefficient is calculated as a projected total number of births divided by the historical total. The coefficients in the table are rounded; calculations were made with unrounded coefficients.
<sup>b</sup> Future numbers of births estimated by multiplying the population growth coefficient with the number of births in Soroka in 2000-2019.
<sup>c</sup> Future preeclampsia cases in Soroka modeled as the number of births in Soroka times the historical preeclampsia incidence ( $5042/128,377 \approx 3.9275\%$ )

### 2.3 Climate projections

Ambient temperature distributions in the region during the historical (2000-2019) and two projected 20-year time intervals (2020-2039 and 2040-2059) were estimated using five climate models under two Shared Socioeconomic Pathways (SSPs): SSP 245 and SSP 585 (Table 2). These SSPs provide a socioeconomic context for future climate change and are characterized by their implications for adaptive and mitigative capacity (Riahi et al., 2017). SSP 245, the middle-of-the-road scenario, has medium mitigative and adaptive capacity. SSP 585 involves continued fossil-fuelled development and low mitigative but high adaptive capacity due to technological development (Riahi et al., 2017).

**Table 2.** Mean temperatures and interval-specific temperature differences predicted by climate models under SSP 245 and 585. From each model, we selected grid points whose latitude and longitude were closest to Beer Sheva

| Model Attributes |  | Mean temperature |  |  | Interval-specific temperature difference <sup>a</sup> |  |
| --- | --- | --- | --- | --- | --- | --- |
| Scenario | Model | 2000-2019 | 2020-2039 | 2040-2059 | 2020-2039 | 2040-2059 |
| SSP 245 | MIROC6 | 24.8 | 25.8 | 26.3 | 1.0 | 1.5 |
|  | MPI-ESM1-2-HR | 18.9 | 19.5 | 19.9 | 0.6 | 1.0 |
|  | MPI-ESM1-2-LR | 20.1 | 20.6 | 21.1 | 0.5 | 1.0 |
|  | MRI-ESM2-0 | 19.8 | 20.6 | 21.0 | 0.9 | 1.2 |
|  | HadGEM3-GC31-LL | 18.5 | 19.4 | 20.5 | 0.9 | 2.0 |
|  | **Scenario mean, <sup>b</sup>** | **20.4** | **21.2** | **21.7** | **0.8** | **1.3** |
| SSP 585 | MIROC6 | 24.8 | 25.9 | 26.7 | 1.1 | 1.9 |
|  | MPI-ESM1-2-HR | 18.9 | 19.3 | 20.3 | 0.4 | 1.4 |
|  | MPI-ESM1-2-LR | 20.1 | 20.8 | 21.6 | 0.7 | 1.5 |
|  | MRI-ESM2-0 | 19.8 | 20.6 | 21.7 | 0.8 | 1.9 |
|  | HadGEM3-GC31-LL | 18.5 | 19.9 | 20.9 | 1.4 | 2.4 |
|  | **Scenario mean, <sup>b</sup>** | **20.4** | **21.3** | **22.3** | **0.9** | **1.8** |

<sup>a</sup> Differences between projected mean temperatures during the future and historical intervals
<sup>b</sup> Average across all models within an SSP scenario
SSP, Shared Socioeconomic Pathway
MIROC6: A climate model developed by a Japanese consortium, designed to simulate the Earth's climate system, including the atmosphere, oceans, and land surface.
MPI-ESM1-2-HR: A high-resolution climate model developed by the Max Planck Institute in Germany, providing detailed simulations of climate processes.
MPI-ESM1-2-LR: A low-resolution climate model developed by the Max Planck Institute in Germany, offering broader but less detailed climate projections than its high-resolution counterpart.
MRI-ESM2-0: A climate model developed by the Meteorological Research Institute in Japan, focusing on interactions between the atmosphere, oceans, and land.
HadGEM3-GC31-LL: A climate model from the UK Met Office's Hadley Centre, used to simulate global climate changes and predict future weather patterns.

The five models were as follows. MIROC6, A climate model developed by a Japanese consortium (Tatebe et al., 2019); MPI-ESM1-2-HR and MPI-ESM1-2-LR: high- and low-resolution climate models, respectively, developed by the Max Planck Institute in Germany (Mauritsen et al., 2019); MRI-ESM2-0: A climate model developed by the Meteorological Research Institute in Japan (Yukimoto et al., 2019); and HadGEM3-GC31-LL: A climate model from the UK Met Office’s Hadley Centre (Andrews et al., 2020). All five climate models are designed to simulate the Earth’s climate system, including the atmosphere, oceans, and land surface. From each model, we selected the grid point geographically closest to Be’er Sheva.

Next, we calculated the differences between the model’s mean temperatures projected for the two future intervals and the historical one. Averaging these differences across all five models within each scenario resulted in four numbers reflecting an expected mean interval-specific temperature increase relative to the mean temperature in 2000-2019 (“interval-specific temperature difference”).

### 2.4 Statistical analysis

Attributable fraction (AF) is generally calculated as:

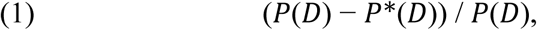

where *P*(*D*) is the disease incidence in the study population, and *P*^∗^(*D*) is the expected incidence in the same population had the exposure been eliminated (Samuelsen and Eide, 2008). With continuous exposures, AF can also be generalized to examine a change in the exposure distribution (the generalized attributable fraction) (Benichou, 2001; Eide and Heuch, 2001). In the context of survival analysis, AF is a function of time, defined as AF(t):

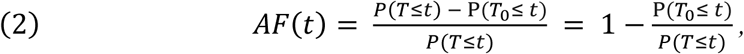

where *T* is the time to event of interest, P(*T*≤ *t*) is the expected probability of an event at or before time *t* in the population (estimated by Kaplan-Meier) given the exposure projected under one of the climate models, and P(*T*_0_ ≤ *t*) is the probability of an event at or before time *t* had the exposure been eliminated/modified for everyone at baseline (Dahlqwist et al., 2016). In this study, P(*T*≤ *t*) is the probability of preeclampsia in the population at or before time *t* under a specific, expected climate change scenario. P(*T_0_* ≤ *t*), on the other hand, represents the probability of the same event under the counterfactual scenario of no climate change, i.e., without future changes in temperature distribution, assuming the temperature distribution in the historical interval of 2000-2019.

Delivery without preeclampsia can be technically described as a competing risk for preeclampsia since once the infant is delivered, preeclampsia diagnosis is no longer possible (Reeder et al., 2023). In this case, AF(t) becomes:

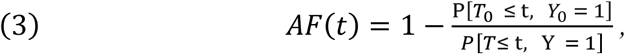

where *T* is the time to preeclampsia or delivery, and *Y* and *Y*_0_ are event variables that equal 1 in the case of the preeclampsia outcome by time *t* and 2 in the case of delivery without preeclampsia by time *t* under a specific climate change scenario or no climate change, respectively (Von Cube et al., 2023). The quantites P[*T* ≤ t, *Y* = 1] and P[*T*_0_ ≤ t, *Y*_0_ = 1] are known as the cumulative incidence functions (CIFs) (Austin et al., 2016).

To estimate an association in the presence of a competing event, a separate (cause-specific) hazard model for an event of interest (here, preeclampsia) was used to calculate cause-specific hazard ratios as implemented in our previous publication (Youssim et al., 2023). Since unlike associations, AF(t) is a function of the CIFs (Von Cube et al., 2023),the adjustment for covariates was implemented using the cause-specific Cox proportional hazard regression model for competing risks from the R package *riskRegression.* This package allows to model both competing events (delivery and preeclampsia) simultaneously and enables extracting the CIF for each event type (Gerds et al., 2015; Therneau, 2022; Therneau et al., 2020). As in the previous study (Youssim et al., 2023), we used the distributed lag models from R package *dlnm* (Gasparrini, 2011) to fit the survival models on weekly exposure histories throughout pregnancy. This was done to mitigate bias arising when examining gestation weeks in separate regression models due to the considerable correlation among temperature exposures during the alternate exposure intervals (Wilson et al., 2017).

The probability P[*T*_0_ ≤ t, *Y*_0_ = 1] was assessed from this initial model by estimating the CIF corresponding to the covariates of each individual in the actual data and averaging it across individuals at each event time (Von Cube et al., 2023). To estimate *P*[*T* ≤ t, *Y* = 1], we firstly added the expected interval-specific differences between the observed and projected temperatures to every value of the original exposure history matrix. This resulted in four exposure distributions: one for each combination of time interval and SSP (Figure 1). The cross-bases of the distributed lag models generated from the exposure matrices were subsequently substituted in the original multi-state model instead of the historical exposure data to obtain CIFs. R package *riskRegression* was used to create a faster algorithm for CIF estimation after fitting a survival model (Gerds et al., 2015).

**Figure 1:**
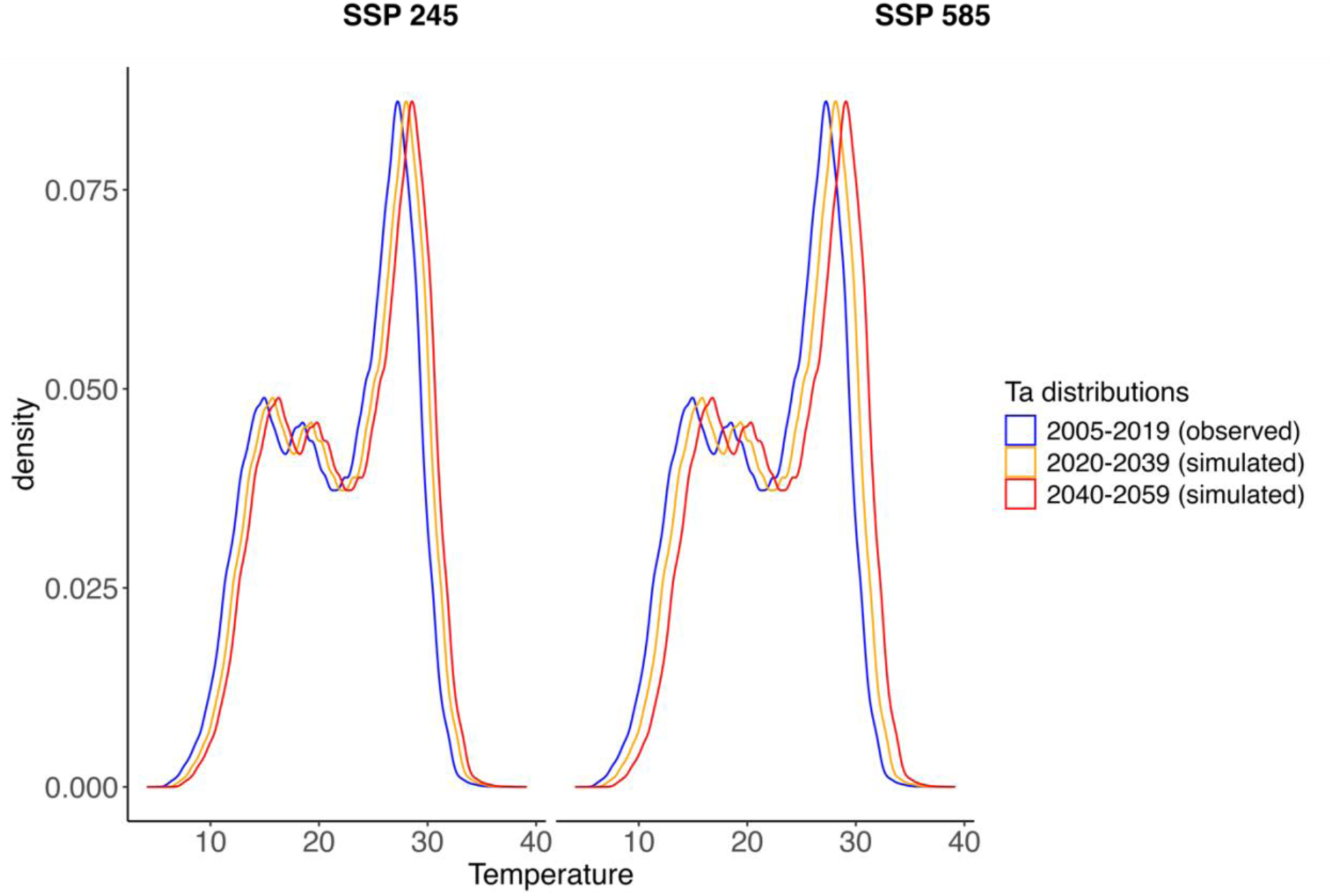
Distributions of temperature observed in 2005-2019 (blue) vs. projected weekly average temperature in 2020-2039 (orange) and 2040-2059 (red). The two latter curves are generated by adding an interval-specific temperature difference predicted under SSP 245 and SSP 585 scenarios to the 2005-2019 temperature distribution. SSP, Shared Socioeconomic Pathway

We used a third-degree polynomial temperature function to facilitate the comparison of different warming scenarios. We subsequently estimated AF(t) by plugging the probabilities P[*T* ≤ *t*, *Y* = 1] and *P*[*T*_0_ ≤ *t*, *Y*_0_ = 1] into equation (3). Next, adjusting the method proposed in Gasparrini and Leone (2014) (Gasparrini and Leone, 2014) and Vicedo-Cabrero et al. (2019) (Vicedo-Cabrera et al., 2019) to survival data, we multiplied the total number of preeclampsia events observed in Soroka in 2005-2019 by the scenario- and interval-specific value of AF(t) at the end of follow-up, obtaining the total attributable numbers of cases for interval and scenario. Annual attributable numbers were calculated assuming identical preeclampsia incidence during 2000-2019 as observed in our study population.

Finally, we projected annual preeclampsia cases in Soroka hospital that consider both demographic and climate change. To do so, the yearly number of preeclampsia cases projected under demographic scenarios for each 20-year interval (Table 1) was multiplied by one plus the annual AF from the corresponding climate scenario. The standard errors of the expected and counterfactual probabilities *P*[*T* ≤ t, *Y* = 1] and *P*[*T*_0_ ≤ t, *Y*_0_ = 1] were estimated using 1,000 bootstrap samples. These standard errors were subsequently employed to calculate the confidence intervals of the AF(t) functions using the delta method (Nevo et al., 2017; Von Cube et al., 2023). All analyses were performed with R version 4.2.1 (R Development Core Team, 2022).

## 3. Results

In total, 247,445 births were registered in Soroka from 2000 to 2019. During 2020-2059, this number is expected to increase by a factor of 1.3 – 2.2, depending on the demographic scenario. Assuming a constant preeclampsia incidence of 3.9% and no climate change, and depending on the time interval and fertility scenario, the modeled annual incidence of preeclampsia during 2020-2059 varies between 646 and 1071 cases, compared to 486 annual cases during 2000-2019 (Table 1).

On average across climate models, according to the SSP 245 scenario, the temperatures in with 2000-2019. Alternatively, if the SSP 585 scenario is realized, the temperatures in 2020-2039 and 2040-2059 are modeled to be 0.9°C and 1.8°C warmer, respectively, compared with 2000-2019 (Table 2). Figure 1 provides a graphical illustration of the shift in the historical temperature distribution had one of the climate scenarios been realized in each of the two intervals.

Figure A.1 visually demonstrates a slight excess in preeclampsia incidence when comparing the cumulative incidence of preeclampsia over the gestation period under the historical and simulated temperature distributions. The excess incidence is strengthened with increasing temperature scenarios. Under the SSP 245 and SSP 585 scenarios, the fractions of preeclampsia attributable to temperature increase in 2020-2039 were close to each other: 2.5% and 2.4% on average over the gestation period, respectively. In comparison, the fractions attributable to temperature increase expected in 2040-2059 under the SSP 245 and SSP 585 scenarios were much higher (3.6% and 4.9% on average during gestation, respectively; Figure 2).

**Figure 2.**
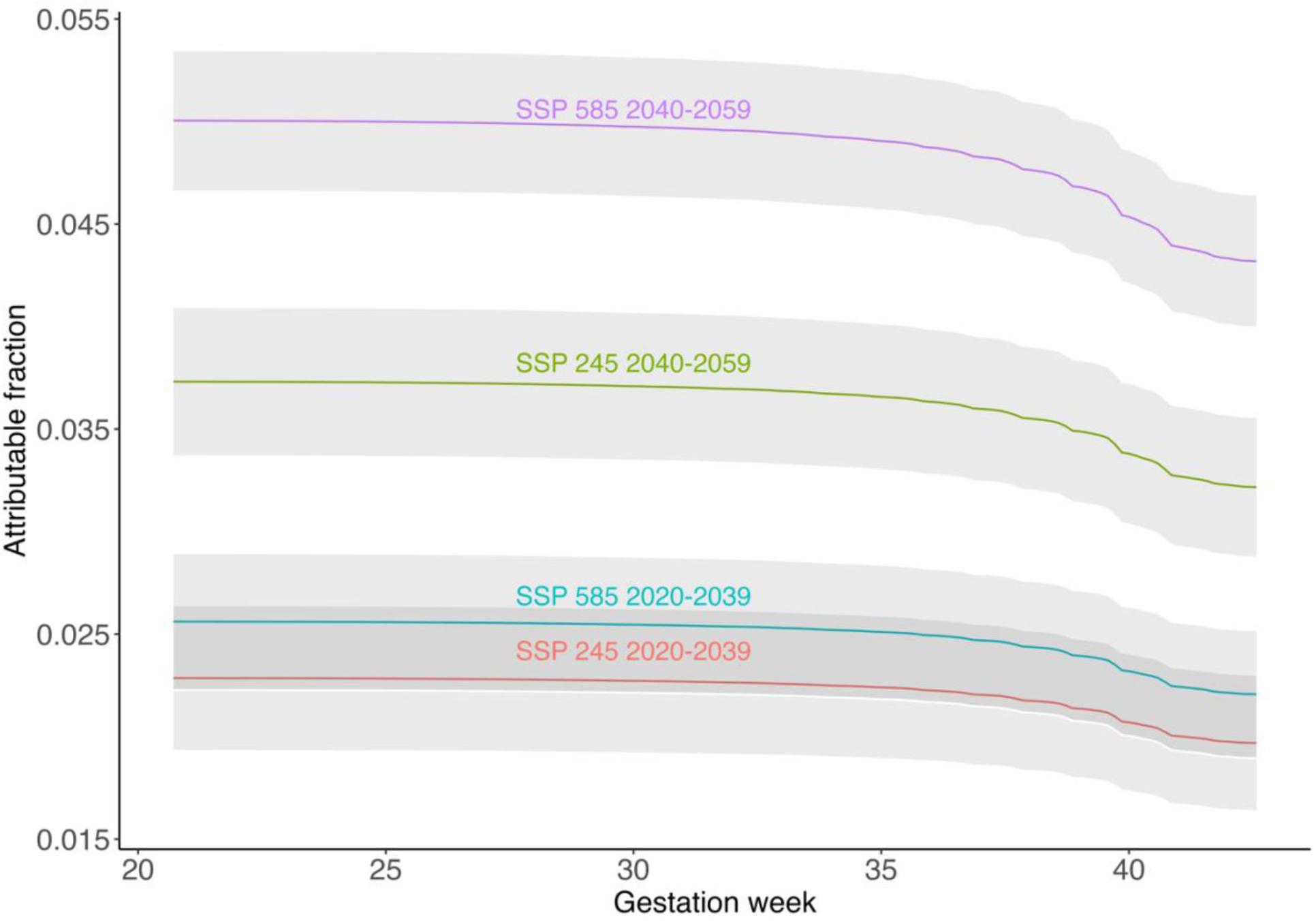
Fractions of preeclampsia attributable to temperature increase as a function of gestation week, by each climate scenario and time interval. Gray bands represent 95% confidence intervals. SSP, Shared Socioeconomic Pathway

Based on these attributable fraction functions and ignoring the demographic changes, had the study population been exposed to warmer temperatures from the beginning of the pregnancy, the estimated number of *excess* annual preeclampsia cases due to temperature changes could range from 10 in 2020-2039 under the SSP 245 scenario to 21 in 2040-2059 under the SSP 585 (Table 3). In annual terms, under the latter scenario, we expect 507 preeclampsia cases in Soroka, representing an increase of 21 cases compared to 486 cases observed during 2000-2019. This increase corresponds to a fraction of 4.3% (95% CI, 4.0% – 4.6%) of excess preeclampsia incidence attributable to the shift of the 2000-2019 temperature distribution by 1.8°C.

**Table 3.** All and temperature-attributable annual preeclampsia cases, by scenario and 20-year interval, before accounting for demographic change.

| Scenario and Interval | Average temperature difference, °C | Average annual preeclampsia cases | Annual cases attributable to temperature increase | Attributable fraction, %<sup>a</sup> | 95% CI, %<sup>b</sup> |
| --- | --- | --- | --- | --- | --- |
| Historical 2000-2019 |  | 486 |  |  |  |
| SSP 245 2020-2039 | 0.8 | 496 | 10 | 2.0 | 1.6, 2.3 |
| SSP 245 2040-2059 | 1.3 | 502 | 16 | 3.2 | 2.9, 3.6 |
| SSP 585 2020-2039 | 0.9 | 497 | 11 | 2.2 | 1.9, 2.5 |
| SSP 585 2040-2059 | 1.8 | 507 | 21 | 4.3 | 4.0, 4.6 |

<sup>a</sup> Attributable fraction functions averaged over the entire pregnancy
<sup>b</sup> Computed using bootstrapping with the delta method
SSP, Shared Socioeconomic Pathway

Finally, Figure 3 presents the projected numbers of preeclampsia cases for the various combinations of fertility and climatic scenarios. Interestingly, while modeled preeclampsia incidence varies by the degree of temperature increase, fertility rates have a larger impact on incidence, with an annual estimate of 1071-1118 cases in the high fertility scenario during 2040-2059, compared with 486 cases during 2000-2019.

**Figure 3.**
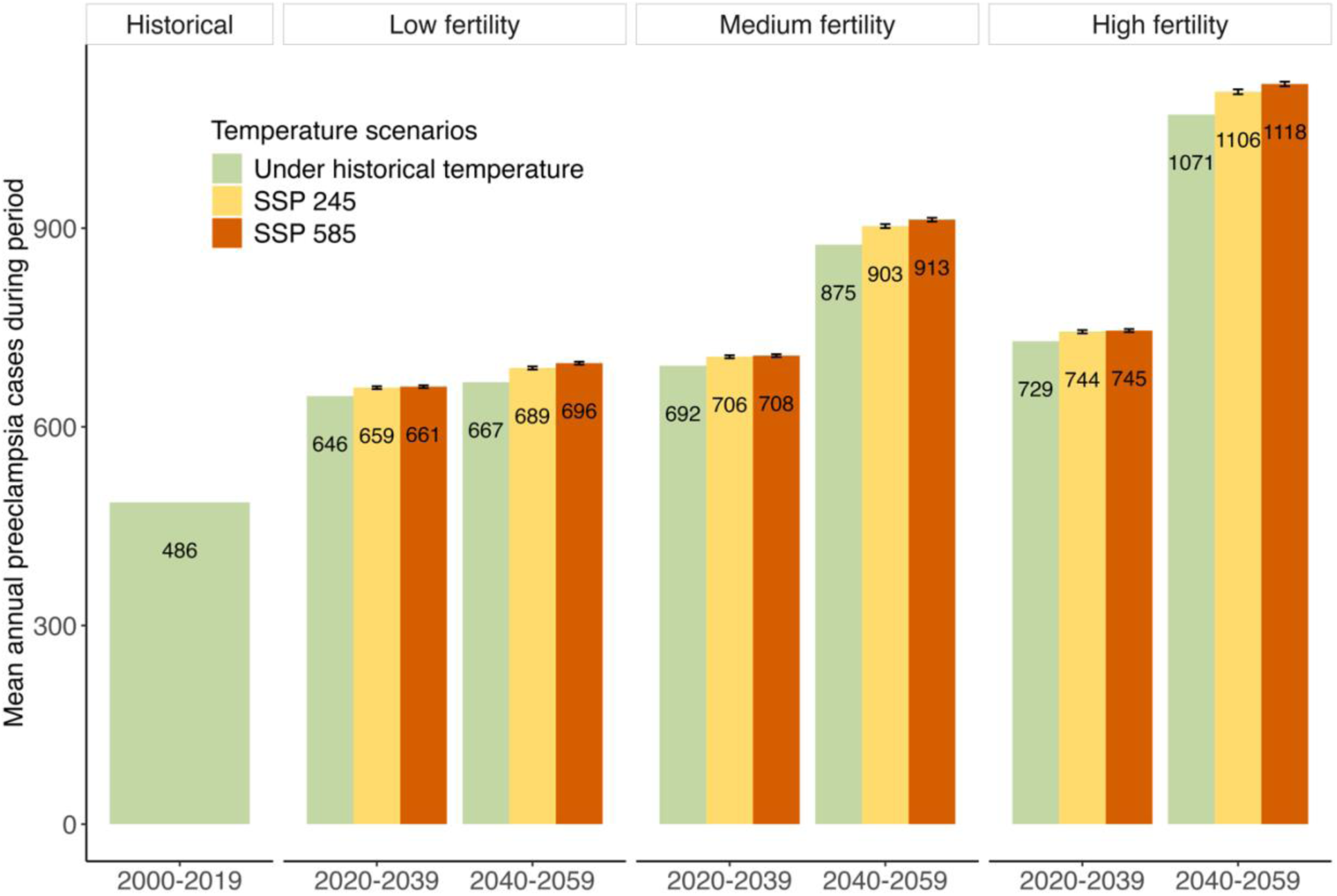
Annual preeclampsia cases are projected under the demographic and climate change scenarios at 20-year intervals. Panels within the figure correspond to demographic scenarios subdivided by 20-year intervals. Green bars show the estimated annual preeclampsia events expected based exclusively on the demographic projections (corresponds to the last column of Table 1). Yellow bars show changes to the demographic projections had the SSP 245 climate scenario been realized. Orange bars show changes to the demographic projections under the SSP 585 climate scenario. Error bars represent 95% confidence intervals. SSP, Shared Socioeconomic Pathway.

## 4. Discussion

In this paper, we modeled future preeclampsia incidence in southern Israel by possible fertility and climatic scenarios for the region. We have based our models on exposure-response relations from our previous work on this population. The current study estimates the attributable fraction of preeclampsia between 2040-2059 due to climate change in the range of approximately ∼3.2-4.3% relative to 2000-2019 temperatures. When adding up the expected increase in the number of births in the region, resulting from the high fertility rates in Israel, our models estimate that this demographic factor will have a much larger impact on the annual number of preeclampsia cases, with ∼30% increase already modeled for 2020-2039 for the low-fertility scenario. Our findings also show that the combined effect of the demographic and climatic factors might reach a stunning 2.3-fold increase in incidence, from 486 to 1118 annual preeclampsia cases when comparing 2000-2019 to 2040-2059, in the more extreme high fertility and climate scenarios.

Given the key role of demographic factors in affecting the public health burden of preeclampsia cases, it is important to understand the evolution of population growth rates in Israel in comparative perspective. While in most affluent, democratic societies, below-replacement fertility has contributed to very low or negative rates of population growth for decades, in Israel, population growth has remained high, fueling increasingly large birth cohorts. In Israel, as in many countries in Africa and other fast-growing areas of the world, the number of babies born each year will continue to increase in the coming decades, spurred in part by population momentum, in the form of large numbers of young people entering the childbearing years. Even if fertility rates were to fall in most population groups in Israel, as per the medium and low fertility scenarios described above, growth in the size of birth cohorts will continue for decades, as a result of population momentum as well as the changing composition of the childbearing population, with increasing proportions in high-fertility subgroups. The implications for the public health burden of preeclampsia cases is apparent from our findings.

Our findings should be interpreted with caution. This study is not meant to provide incidence forecasts or predictions. Instead, this is a relatively simple mathematical modeling of the potential effects of two factors - fertility and climate - on future preeclampsia incidence under several strong assumptions. Except for the usual statistical assumptions, typical of the statistical tools we used, our models further assume causality of the temperature-preeclampsia relations and a constant incidence proportion (3.9%) given the historical temperature distribution. The latter assumption ignores possible changes in risk factors and treatment options for preeclampsia, which are both unpredictable. For example, our data show a reduction in preeclampsia incidence after 2010, probably due to the implementation of prophylactic aspirin treatment in susceptible women (Than et al., 2023). However, for unknown reasons, preeclampsia incidence was raised back to its baseline after 2016, showing that many unpredictable factors probably impact incidence.

Another assumption of our modeling method is that of no adaptation to the changing climate with regard to its effect on preeclampsia. This assumption is also not testable with existing data, which are not rich enough and do not span long. Only future studies can shed light on its plausibility. Physiological and technological adaptations are undoubtedly possible, and there is some evidence for adaptation to the effect of temperatures on mortality (Huber et al., 2022; Wang et al., 2018).

In addition, one should note that all components of our model have their uncertainties (Vanos et al., 2020). While the uncertainty in the exposure-response function of our epidemiological model is quantified as part of that model in traditional methods, the uncertainty in climate scenarios is represented by averaging over five global climate models, and some of the uncertainty in fertility scenarios is defined by comparing the three demographic projections. This further emphasizes that our results present a simple modeling approach to the effects on future incidence and not a forecast. Still, we believe that modeling potential scenarios is valuable in improving the planning of future health services, a task that is always done under high uncertainty.

We used local epidemiological, climatic, and demographic data and models to model incidence in a specific region. A recent review found that while projecting the heat health burden is a growing area of research, some regions remain understudied, including the Middle East, and morbidity is rarely explored, with most studies focusing on mortality (Cole et al., 2023). This study contributes to bridging both of these two gaps in the literature. This geographic focus of this study improves the validity of our results but also limits their generalizability. However, our study population is very diverse in terms of infrastructure and household environments (Shashar et al., 2020). Our methodology, based on the principles defined by Vicedo-Carbera et al. (Vicedo-Cabrera et al., 2019) and applying them to survival analyses, can also be used to develop similar models for other regions. Moreover, the approach developed in this paper can be applied to project the incidence of other intermediate-term clinical endpoints, even beyond adverse pregnancy outcomes. Different mixes of demographic, climatic, and epidemiological models for specific regions and endpoints may produce different results than ours.

In summary, modeling disease incidence is crucial for planning health services for the population. Such planning should be done well in advance since planning and building new medical units and hospitals may take decades. Moreover, preeclampsia is just one example of a disorder that may be impacted by climate change. Even in the context of pregnancy, strong associations with ambient temperature were shown with much more prevalent complications, such as preterm birth. Accordingly, regional and national health authorities should plan their health services considering demographic and climate change projections.

## Abbreviations

AF attributable fraction

CI 95% confidence interval

ICBS Israel Central Bureau of Statistics

SSP Shared Socioeconomic Pathway

## Ethical approval

The Soroka institutional review board approved the study, including the exemption from informed consent due to the sample size and deidentified nature of the records.

## Data availability

The authors do not have permission to share data.

## Appendix A

**Figure A.1:**
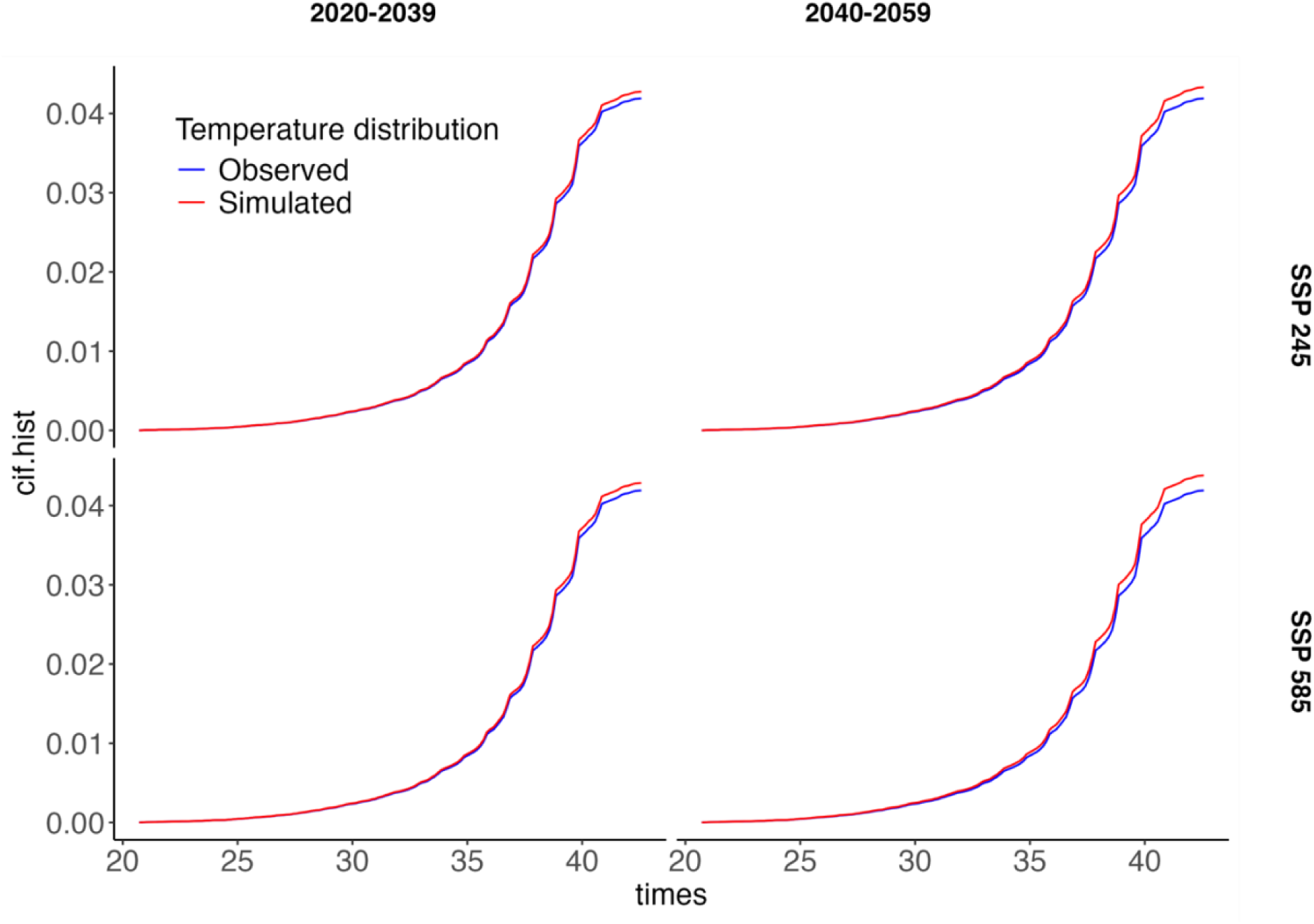
Cumulative incidence functions (CIFs) for observed and projected weekly temperature, by SSP scenario and time interval. CIFs predicted by the multi-state with distributed-lag Cox models using the historical temperature distribution (blue curves) and counterfactual temperature distributions (red curves) based on two climate scenarios and two future intervals. SSP, Shared Socioeconomic Pathway

